# Anti-Spike, anti-Nucleocapsid and neutralizing antibodies in SARS-CoV-2 inpatients and asymptomatic carriers

**DOI:** 10.1101/2020.05.12.20098236

**Authors:** Etienne Brochot, Baptiste Demey, Antoine Touzé, Sandrine Belouzard, Jean Dubuisson, Jean-Luc Schmit, Gilles Duverlie, Catherine Francois, Sandrine Castelain, Francois Helle

## Abstract

**Objective:** The objective of this study was to monitor the anti-SARS-CoV-2 antibody response in infected patients.

**Methods:** In order to assess the time of seroconversion, we used 151 samples from 30 COVID-19 inpatients and monitored the detection kinetics of anti-S1, anti-S2, anti-RBD and anti-N antibodies with in-house ELISAs. We also monitored the presence of neutralizing antibodies in these samples as well as 25 asymptomatic carrier samples using retroviral particles pseudotyped with the spike of the SARS-CoV-2.

**Results:** We observed that specific antibodies were detectable in all inpatients two weeks post-symptom onset. The detection of the SARS-CoV-2 Nucleocapsid and RBD was more sensitive than the detection of the S1 or S2 subunits. Neutralizing antibodies reached a plateau two weeks post-symptom onset and then declined in the majority of inpatients. Furthermore, neutralizing antibodies were undetectable in 56% of asymptomatic carriers.

**Conclusions:** Our results raise questions concerning the role played by neutralizing antibodies in COVID-19 cure and protection against secondary infection. They also suggest that induction of neutralizing antibodies is not the only strategy to adopt for the development of a vaccine. Finally, they imply that anti-SARS-CoV-2 neutralizing antibodies should be titrated to optimize convalescent plasma therapy.

**Highlights:**

- Specific antibodies are detectable in 100% COVID-19 inpatients two weeks post-symptom onset.
- The detection of the SARS-CoV-2 Nucleocapsid and Receptor Binding Domain is more sensitive than the detection of the S1 or S2 subunits.
- Neutralizing antibodies reach a plateau two weeks post-symptom onset and then decline in the majority of inpatients.
- Neutralizing antibodies are undetectable in the majority of asymptomatic carriers.

## Background

The Severe Acute Respiratory Syndrome Coronavirus 2 (SARS-CoV-2) has recently emerged and caused a human pandemic of coronavirus disease 2019 (COVID-19).^1–3^ Among the coronaviruses structural proteins, the Spike (S) and the Nucleocapsid (N) proteins are the main immunogens.^4^ The S protein consists of two subunits, S1 which contains the Receptor Binding Domain (RBD) and S2. The kinetics of antibody detection is essential for the selection of commercial serological assays and the interpretation of the results. Some manufacturers have decided to target the S1 and/or S2 subunits whereas others chose the RBD or the N protein. Furthermore, the neutralizing antibody (NAb) response against the SARS-CoV-2 remains poorly documented and it is still unknown how long cured patients will be protected against new infection.^5,6^ The objective of our study was thus to monitor the anti-SARS-CoV-2 antibody response in infected patients.

## Study design

### Study population and specimen

Thirty patients diagnosed SARS-CoV-2 positive by RT-PCR on a nasopharyngeal swab sample, between 25 February and 23 March 2020 at the Amiens University Medical Center, were enrolled in the study. The general information was extracted from electronic medical records and the clinical characteristics of the 30 inpatients are described in Supplementary Table 1. Inpatients were considered as having mild disease when needing non-intensive care or severe disease when needing intensive care. Samples from patients diagnosed positive for other human coronaviruses (OC43 (n=5), 229E (n=4), NL63 (n=2) or HKU1 (n=1)) were also tested (Supplementary Table 2). Finally, we also used samples from 25 asymptomatic carriers (Supplementary Table 3) that were diagnosed SARS-CoV-2 positive using commercial serological tests (LIAISON® SARS-CoV-2 IgG from DiaSorin and/or ELISA SARS-CoV-2 (IgG) from EUROIMMUN). All plasmas were decomplemented at 56°C for 30 min. The study was approved by the institutional review board of the Amiens University Medical Center (number PI2020_843_0046, 21 April 2020).

### In-house ELISAs

MaxiSorp Nunc-immuno 96-well plates were coated with a 1 μg/mL solution of SARS-CoV-2 S1, S2, RBD or N antigen (The Native Antigen Company, United Kingdom), overnight at 4°C. Wells were blocked with 1% fetal bovine serum for 1 hour at 37°C. Then, 100 µL of diluted plasmas (1:100 for S1, S2 and RBD or 1:200 for N) were added and incubated for 1 h at 37°C. After washing 4 times, plates were incubated with peroxydase conjugated mouse anti-human IgG (Southern Biotech, 1/6000). After 4 washes, 100 µL of o-phenylenediamine peroxidase substrate was added at room temperature in the dark. The reaction was stopped with H_2_SO_4_ solution 15 min later. The optical density was measured at 490 nm. All samples were run in triplicate. To establish the specificity of each assay, 40 pre-pandemic sera from 2019 were tested. Each cut-off values were defined as the means plus 3 standard deviations obtained with these samples.

### Neutralization assay

Retroviral particles pseudotyped with the S glycoprotein of the SARS-CoV-2 (SARS-CoV-2pp) were produced as described previously ^7^, with a plasmid encoding a human codon-optimized sequence of the SARS-CoV-2 spike glycoprotein (accession number: MN908947). Supernatants containing the pseudotyped particles were harvested at 48, 72 and 96 h after transfection, pooled and filtered through 0.45-µm pore-sized membranes. Neutralization assays were performed by preincubating SARS-CoV-2pp and diluted plasma for 1 h at room temperature before contact with Vero cells (ATCC® CCL-81™) that were transiently transfected with the plasmids pcDNA3.1-hACE2 and pcDNA3.1-TMPRSS2 48 h before inoculation. Luciferase activities were measured 72 h post-infection, as indicated by the manufacturer (Promega). The NAb titers were defined as the highest dilution of plasma which resulted in a 90% decrease of the infectivity. Retroviral particles pseudotyped with the G glycoprotein of the Vesicular Stomatitis Virus (VSVpp) were used to control the specificity of the neutralization.

### Statistical analysis

Quantitative variables were expressed as the median and compared using Student’s t-test. The Pearson correlation coefficient was used to measure the strength of a linear association between two quantitative variables. Statistical analyses were performed using GraphPad Prism 5. A two-sided P-value < 0.05 was considered statistically significant.

## Results

### Antibody response in SARS-CoV-2 infected inpatients

In order to accurately assess the time of seroconversion, we used 151 samples from 30 patients hospitalized at the Amiens University Medical Center for a COVID-19 (see Supplementary Table 1) and monitored the kinetics of detection of anti-S1, anti-S2, anti-RBD and anti-N antibodies with in-house ELISAs. Importantly, plasmas from twelve patients that had previously been infected with other coronaviruses (OC43 (n=5), 229E (n=4), NL63 (n=2) or HKU1 (n=1)) showed minimal cross-reactivity, which highlights the specificity of these assays (Supplementary Table 2). We observed that antibodies targeting the N protein and the RBD were the earliest to be detected (Fig. 1A). Thirteen days post-symptom onset, 100% of inpatients had detectable antibodies to both proteins. A similar profile was observed for anti-S2 antibodies but with a mean time lag of two days. Antibodies to the S1 subunit were the last to be detected and remained undetectable for two inpatients. High levels of anti-N and anti-RBD antibodies were detected in the large majority of samples obtained fourteen days post-symptom onset whereas very heterogeneous levels of anti-S1 antibodies were found in the same samples (Fig. 1B). The correlations between each ELISA are shown in Supplementary Fig. 1 and clearly demonstrate that detection of the N protein and/or the RBD is more sensitive than the detection of the S1 (Supplementary Fig. 1B and 1C) or the S2 subunit (see Supplementary Fig. 1D and 1E). Significant differences were observed between mild disease versus severe disease patients for anti-S1, anti-S2 and anti-N antibody levels, from eight days post-symptom onset (Supplementary Fig. 2A). A slight difference was observed for anti-N antibody levels according to the sex, from fourteen days post-symptom onset (Supplementary Fig. 2B). Finally, a significant difference was observed for anti-S1 and anti-S2 antibodies according to the age, between eight and fourteen days post-symptom onset (Supplementary Fig. 2C).

**Fig. 1.**
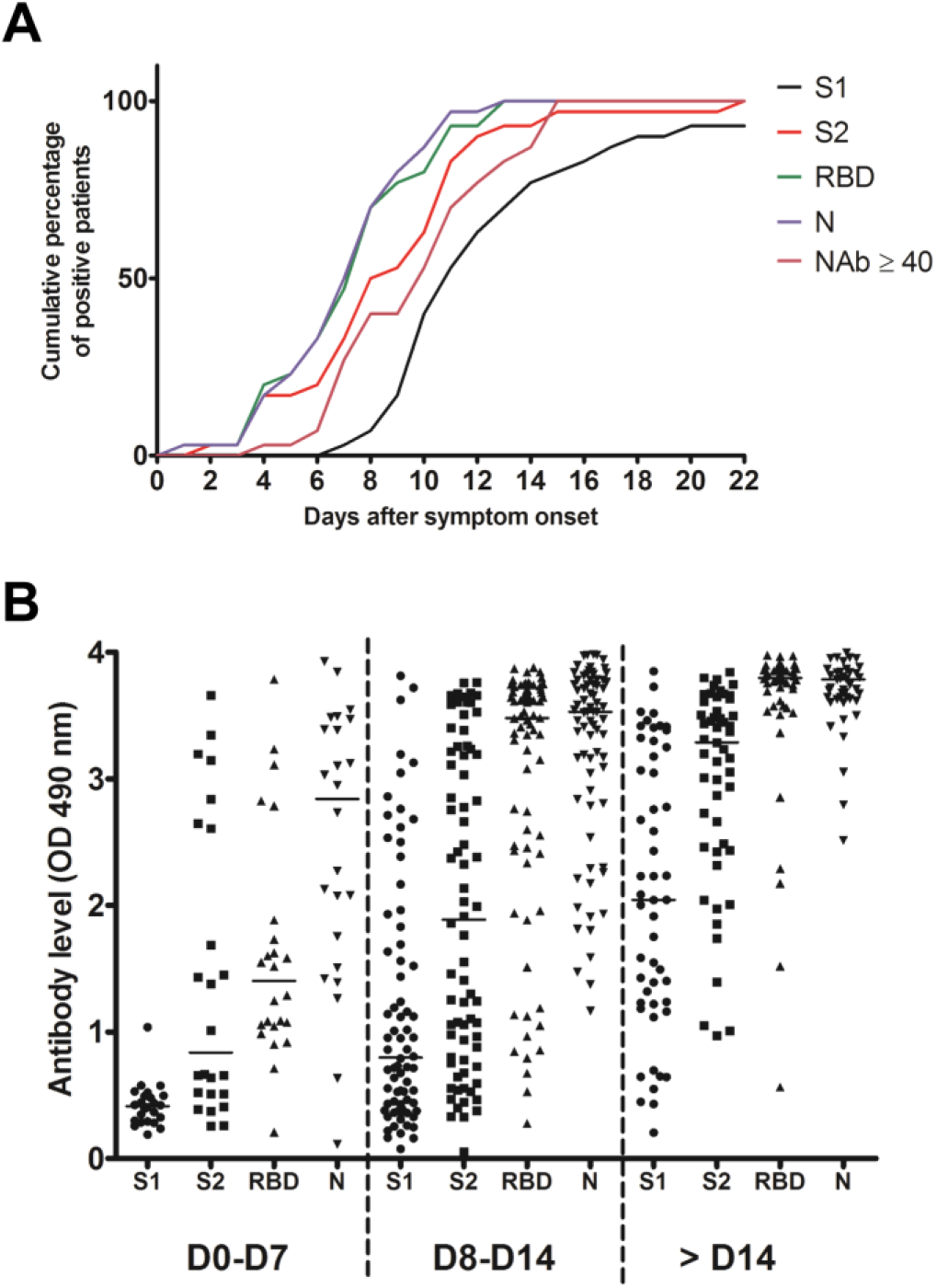
Antibody response in SARS-CoV-2 infected inpatients. (A) Kinetics of anti-S1, anti-S2, anti-RBD, anti-N and NAb detection in 30 COVID-19 inpatients post-symptom onset. (B) Evolution of the anti-S1, anti-S2, anti-RBD and anti-N antibody levels during the first month post-symptom onset.

### NAb response to SARS-CoV-2 in COVID-19 inpatients

We also monitored the presence of NAbs in all plasma samples using SARS-CoV-2pp.^7^ Importantly, Ni *et al*. recently demonstrated that there was a significantly positive correlation in the NAb titers between such pseudotyped particles and the native SARS-CoV-2.^8^ The results obtained for each inpatient are presented in Fig. 2. One sample of each inpatient was also used to perform dose-response curves with VSVpp and no inhibition was observed, demonstrating that the neutralization observed with the COVID-19 inpatient plasmas was specific to the SARS-CoV-2 (Supplementary Fig. 3A). Furthermore, plasmas from the twelve patients that had previously been infected with other coronaviruses did not have any effect on SARS-CoV-2 pseudotype infectivity (Supplementary Table 2 and Supplementary Fig. 3B). As expected, our results demonstrate that the NAb production kinetic correlates with the production of antibodies targeting the S1, S2 subunits as well as the RBD and we detected NAbs in all COVID-19 inpatients fifteen days post-symptom onset (Fig. 1A). The NAb titers increased from one week post-symptom onset and reached a plateau one week after (Fig. 3). However, the NAb titers reached were variable between inpatients, 17% generated low levels of NAbs (40 ≤ titers < 160), 73% intermediate levels (160 ≤ titers < 1280) and 10% high levels (1280 ≤ titers) (Fig. 3A). We also had the opportunity to monitor the presence of NAbs in late samples of eleven inpatients (≥40 days post-symptom onset) and we observed that the NAb titer dropped to low or undetectable level in most of these samples (Fig. 3B). Significantly higher NAb titers were observed in inpatients with severe forms (p=0.04; Supplementary Fig. 4A) and in women (p=0.03; Supplementary Fig. 4B) from 14 days post-symptom onset. In contrast, no significant difference was observed according to the age (Supplementary Fig. 4C). Furthermore, we found poor correlations between NAb titers and anti-S1 (r=0.4573), anti-S2 (r=0.3852), anti-N (r=0.3629) or anti-RBD (r=0.3277) antibody levels, as well as white blood cells (r=0.2384) and lymphocytes counts (r=0.3696) (Fig. 4).

**Fig. 2.**
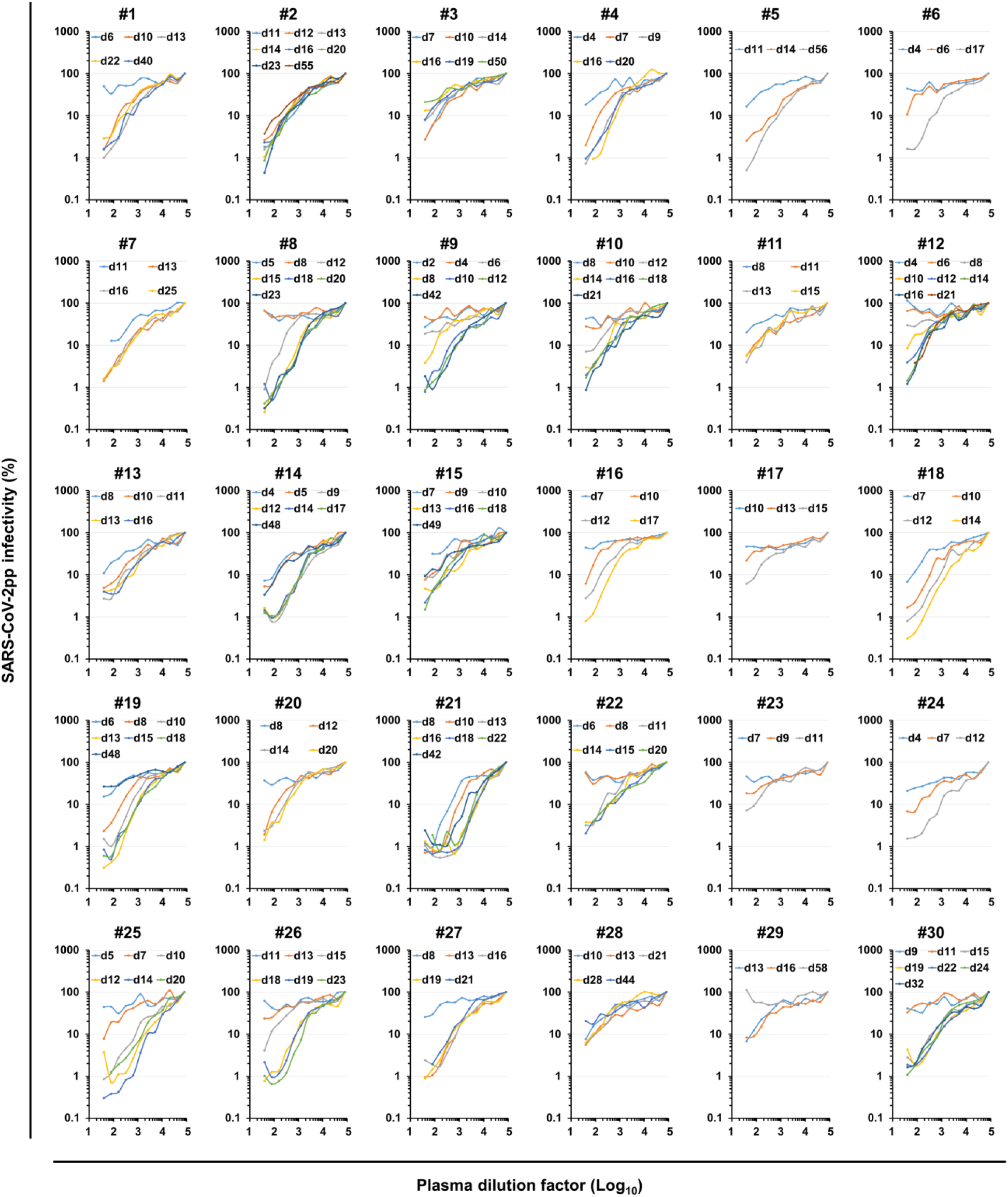
NAb response to SARS-CoV-2 in COVID-19 inpatients. SARS-CoV-2pp were preincubated with serially diluted plasma obtained from 30 COVID-19 inpatients (#1 to #30) at different days post-symptom onset (d2 to d58). Dose response curves represent the means of normalized infectivity (%) from two independent experiments performed in duplicate. Error bars have been omitted for clarity.

**Fig. 3.**
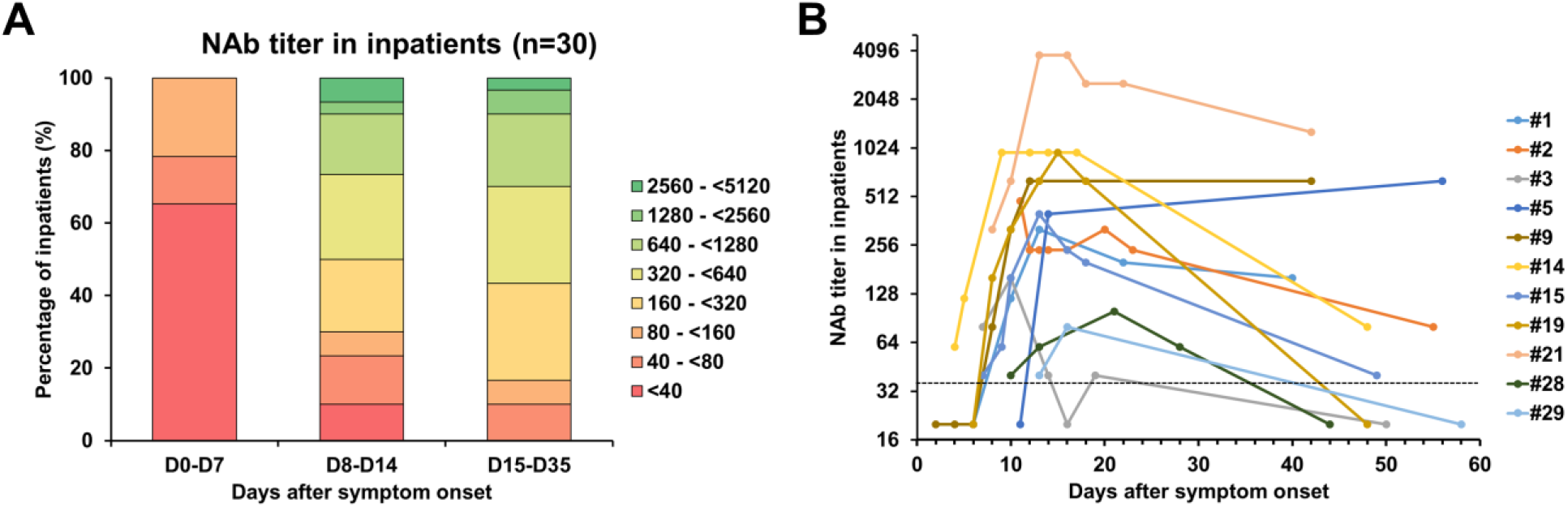
(A) Evolution of the NAb titer in 30 COVID-19 inpatients during the first month post-symptom onset. (B) Evolution of the NAb titer in 11 COVID-19 inpatients after more than 40 days post-symptom onset. The dashed line indicates the cut-off of the assay.

**Fig. 4.**
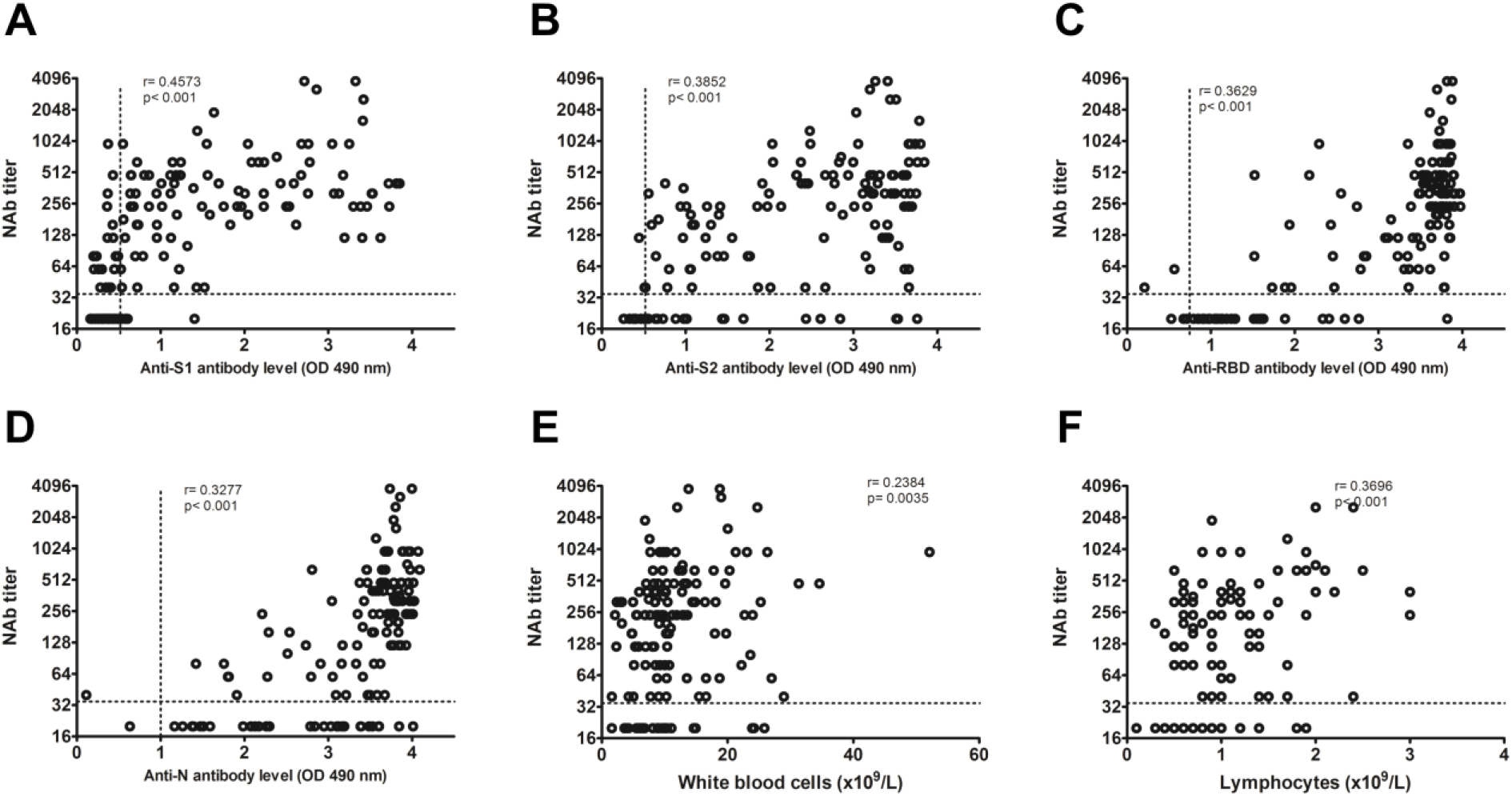
Correlations between NAb titers and anti-S1, anti-S2, anti-RBD and anti-N antibody levels as well as white blood cells counts or lymphocyte counts. (A) NAb titer versus anti-S1. (B) NAb titer versus anti-S2. (C) NAb titer versus anti-RBD. (D) NAb titer versus anti-N. (E) NAb titer versus white blood cells counts. (F) NAb titer versus lymphocyte counts. Dashed lines indicate assay cutoffs for positivity. OD, optical density.

### SARS-CoV-2 NAbs in asymptomatic carrier samples

Finally, we monitored the presence of NAbs in plasma samples from 25 asymptomatic carriers. It is important to note that we could not establish when these patients had been infected since they were asymptomatic but it probably occurred more than one week before sampling since they were confirmed seropositive using commercial serological assays as well as in-house ELISAs (Supplementary Table 3). As shown in Fig. 5, NAbs were below the detection limit of our assay in the majority of these plasma samples (56%, 14/25). Low NAb levels (40 ≤ titers < 160) were found in 28% of these patients (7/25). Three patients had intermediate NAb levels (160 ≤ titers < 1280) and only one showed a high NAb titer (≥1280).

**Fig. 5.**
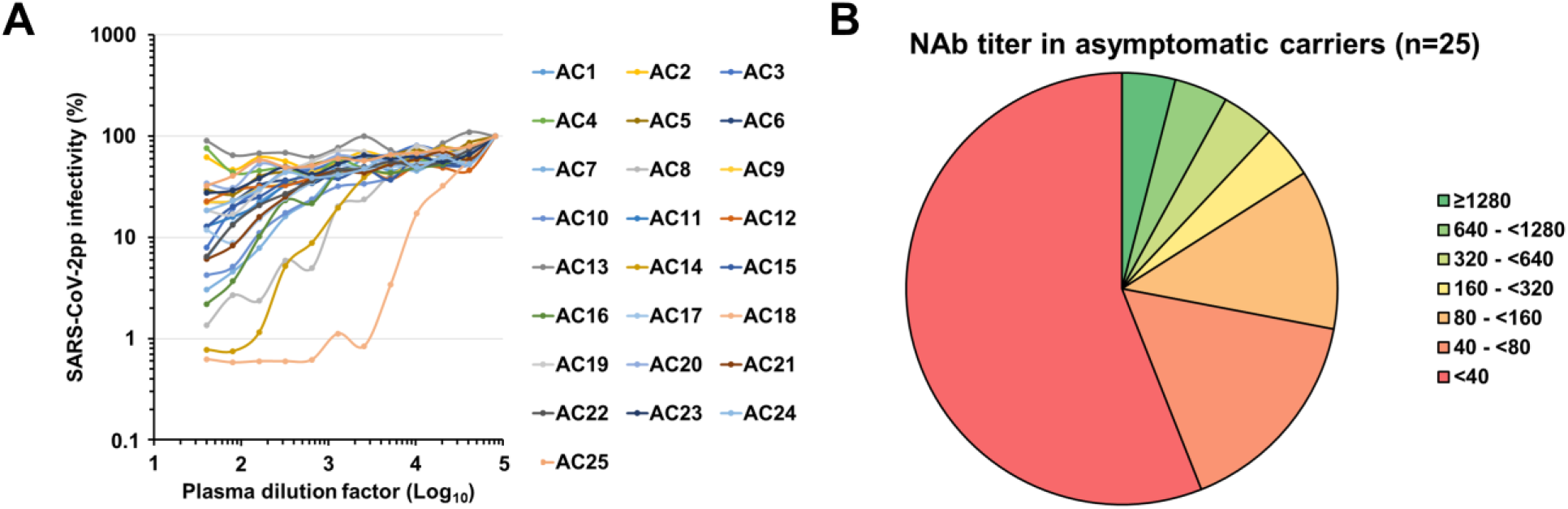
SARS-CoV-2 NAbs in asymptomatic carrier samples. (A) SARS-CoV-2pp were preincubated with serially diluted plasma obtained from 25 asymptomatic carriers that were confirmed with commercial serological assays (AC1 to AC25). Dose response curves represent the means of normalized infectivity (%) from two independent experiments performed in duplicate. Error bars have been omitted for clarity. (B) Determination of the NAb titer in plasma samples from 25 asymptomatic carriers.

## Discussion

Commercial serological assays that are complementary to direct viral detection of the SARS-CoV-2 by RT-PCR have recently become available but they need to be finely evaluated.^9,10^ We only tested IgG detection since recent data showed that anti-SARS-CoV-2 IgG levels increase at the same time or earlier than IgM levels.^11^ With our four in-house ELISAs, we showed that the detection of the RBD and the N protein may be more suitable since it was highly or slightly more sensitive than the detection of S1 or S2, respectively. The presence of cryptic epitopes in the RBD could explain why in some cases anti-RBD antibodies are detected whereas anti-S1 antibodies are not.^12^

As other groups, we also report that COVID-19 patients generate variable levels of NAbs that reach a plateau two weeks post-symptom onset without correlation with clinical courses. ^8,13–16^ This raises questions concerning the role played by NAbs and suggest that other arms of the immune system may play a more important role in COVID-19 cure. Our results also indicate that the SARS-CoV-2 does not induce a prolonged neutralizing antibody response since we observed a drop of the NAb titer for several patients a few weeks after infection, which has also been observed by Wang *et al*. on a small cohort.^14^ Furthermore, we observed that most of asymptomatic carriers do not generate NAbs. The longevity of the protection against reinfection is thus questionable. However, the immunological memory would certainly protect against severe disease if reinfection would occur. These results also suggest that induction of NAb production is not the only strategy to adopt for the development of a SARS-CoV-2 vaccine. Finally, since we observed poor correlations between NAb titers and S1, S2, RBD or N binding, our results imply that anti-SARS-CoV-2 NAbs should be titrated to optimize convalescent plasma therapy.^17,18^

## Data Availability

Most of the data generated or analysed during this study are included in this published article (and its supplementary information files). Additional supporting data are available from the corresponding authors on request.

## Acknowledgments

We thank the study participants who donated their blood samples for this project, and the healthcare workers who care for COVID-19 patients. This work was supported by Flash Covid-19 funds from the Amiens University Medical Center and the French “Agence Nationale de la Recherche”.

---

**Supplementary Table 1.**
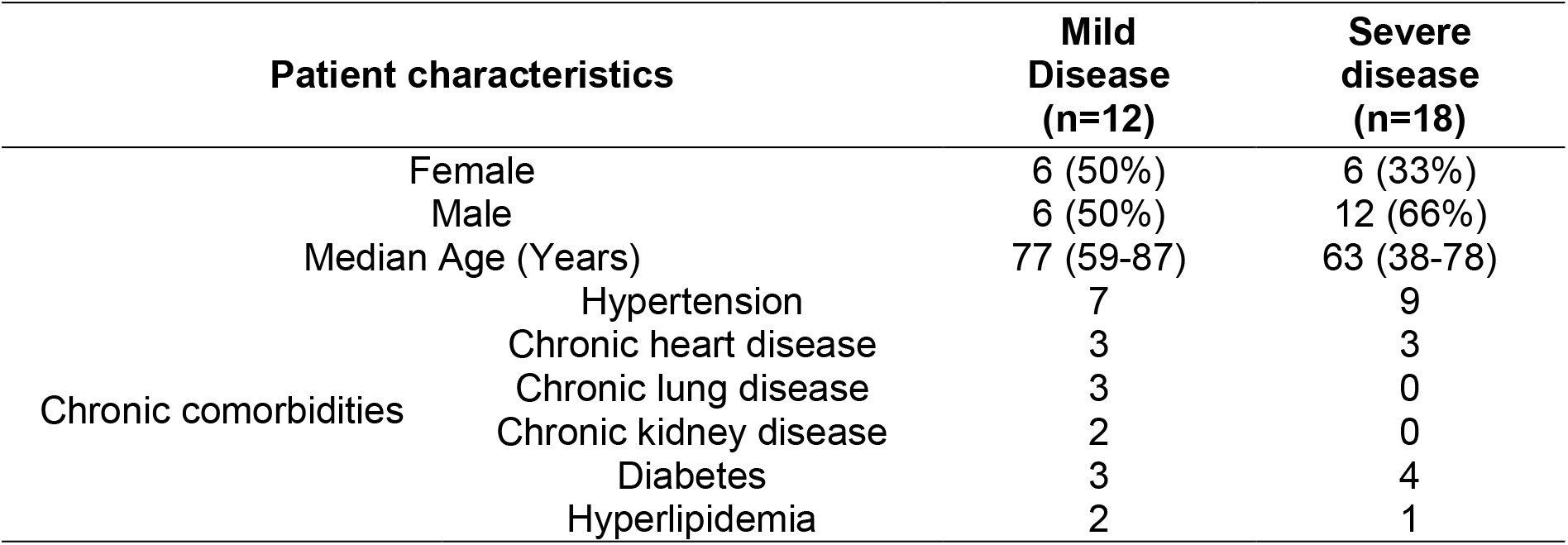
Characteristics of the 30 inpatients included in the study

**Supplementary Table 2.** Patients previously infected with endemic coronaviruses used as control in the study

| Patient | Coronavirus strain | Days post-diagnostic | Detection of IgG in our in house ELISA assays |  |  |  | SARS-CoV-2 NAb titer |
| --- | --- | --- | --- | --- | --- | --- | --- |
|  |  |  | S1 | S2 | RBD | NP |  |
| C1 | OC43 | 174 | - | - | +/- | - | <40 |
| C2 | 229E | 39 | - | - | - | - | <40 |
| C3 | NL63 | 561 | - | - | - | - | <40 |
| C4 | OC43 | 170 | - | - | - | - | <40 |
| C5 | OC43 | 176 | - | - | - | - | <40 |
| C6 | NL63 | 16 | - | - | - | - | <40 |
| C7 | 229E | 14 | - | - | + | + | <40 |
| C8 | HKU1 | 24 | - | - | - | - | <40 |
| C9 | OC43 | 301 | +/- | - | - | +/- | <40 |
| C10 | 229E | 56 | - | - | - | - | <40 |
| C11 | 229E | 761 | - | - | - | +/- | <40 |
| C12 | OC43 | 86 | - | - | - | - | <40 |

+, positive ; -, negative ; +/-, equivocal

**Supplementary Table 3.** SARS-CoV-2 asymptomatic carriers included in the study

| Patient | Detection of IgG in<br>in house ELISAs |  |  | Diasorin<br>assay | Euro-<br>immun<br>assay | SARS-CoV-2 NAb titer |
| --- | --- | --- | --- | --- | --- | --- |
|  | S1 | S2 | NP |  |  |  |
| AC1 | + | + | + | + | +/- | 40 |
| AC2 | + | +/- | + | - | + | <40 |
| AC3 | + | + | + | + | + | 40 |
| AC4 | +/- | + | + | + | - | <40 |
| AC5 | - | + | + | + | - | <40 |
| AC6 | +/- | + | + | + | +/- | <40 |
| AC7 | + | + | + | + | + | 160 |
| AC8 | + | + | + | + | + | 640 |
| AC9 | + | + | + | + | + | <40 |
| AC10 | + | + | + | + | + | 80 |
| AC11 | + | + | + | - | + | <40 |
| AC12 | - | + | + | + | - | <40 |
| AC13 | + | +/- | + | - | + | <40 |
| AC14 | + | + | + | + | + | 480 |
| AC15 | + | + | + | +/- | + | <40 |
| AC16 | + | + | + | + | + | 120 |
| AC17 | + | + | + | + | + | 80 |
| AC18 | + | + | + | + | + | 5120 |
| AC19 | + | + | + | + | + | <40 |
| AC20 | + | + | + | + | + | <40 |
| AC21 | + | + | + | + | + | 60 |
| AC22 | + | + | + | + | + | 60 |
| AC23 | + | + | + | + | + | <40 |
| AC24 | + | + | + | + | + | <40 |
| AC25 | + | + | + | + | + | <40 |

+, positive ; -, negative ; +/-, equivocal

**Supplementary Fig. 1.**
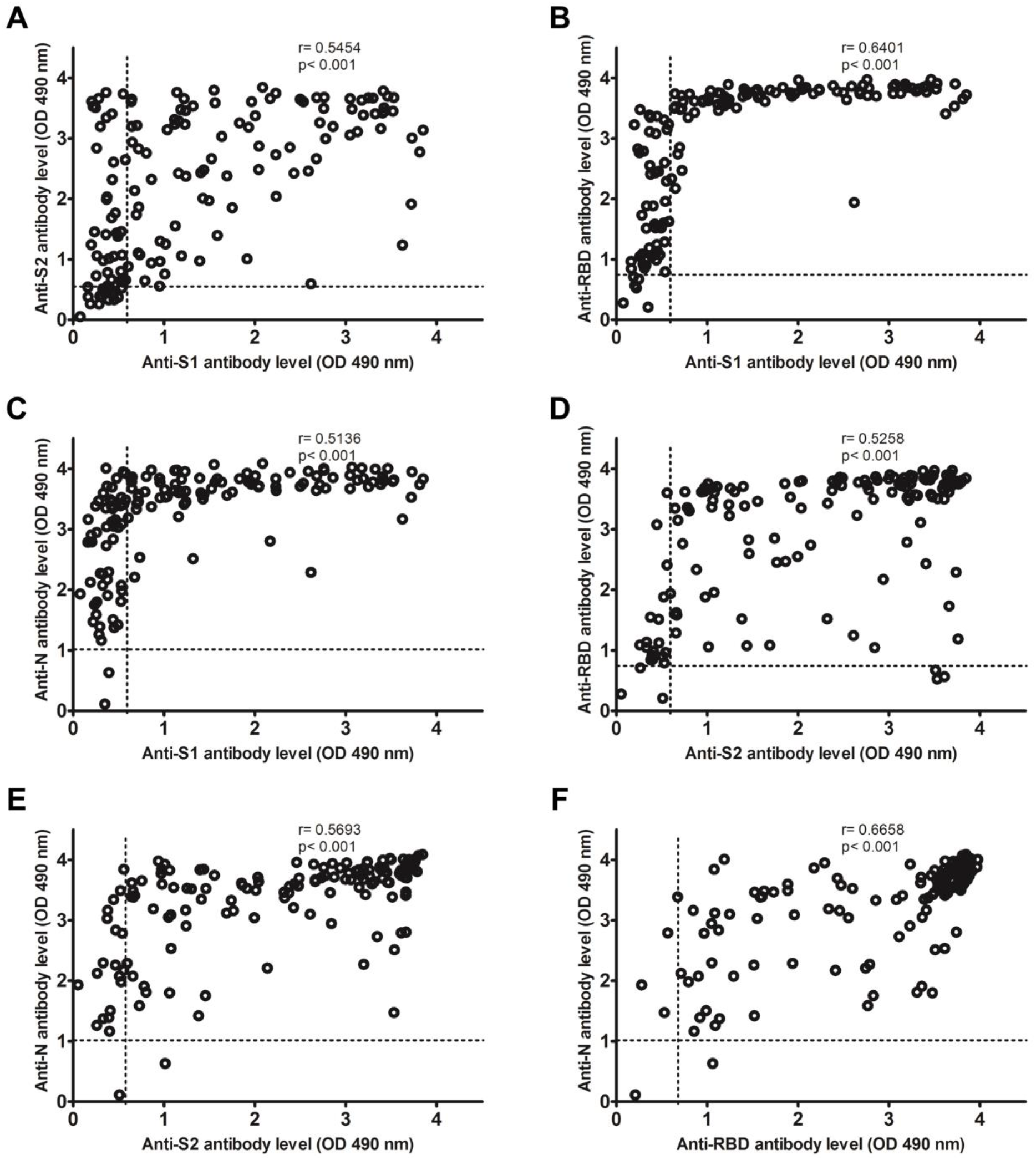
Correlations between anti-S1, anti-S2, anti-RBD and anti-N levels in inhouse ELISAs. (A) anti-S1 versus anti-S2. (B) anti-S1 versus anti-RBD. (C) anti-S1 versus anti-N. (D) anti-S2 versus anti-RBD. (E) anti-S2 versus anti-N. (F) anti-RBD versus anti-N. Dashed lines indicate assay cut-offs for positivity. OD, optical density.

**Supplementary Fig. 2.**
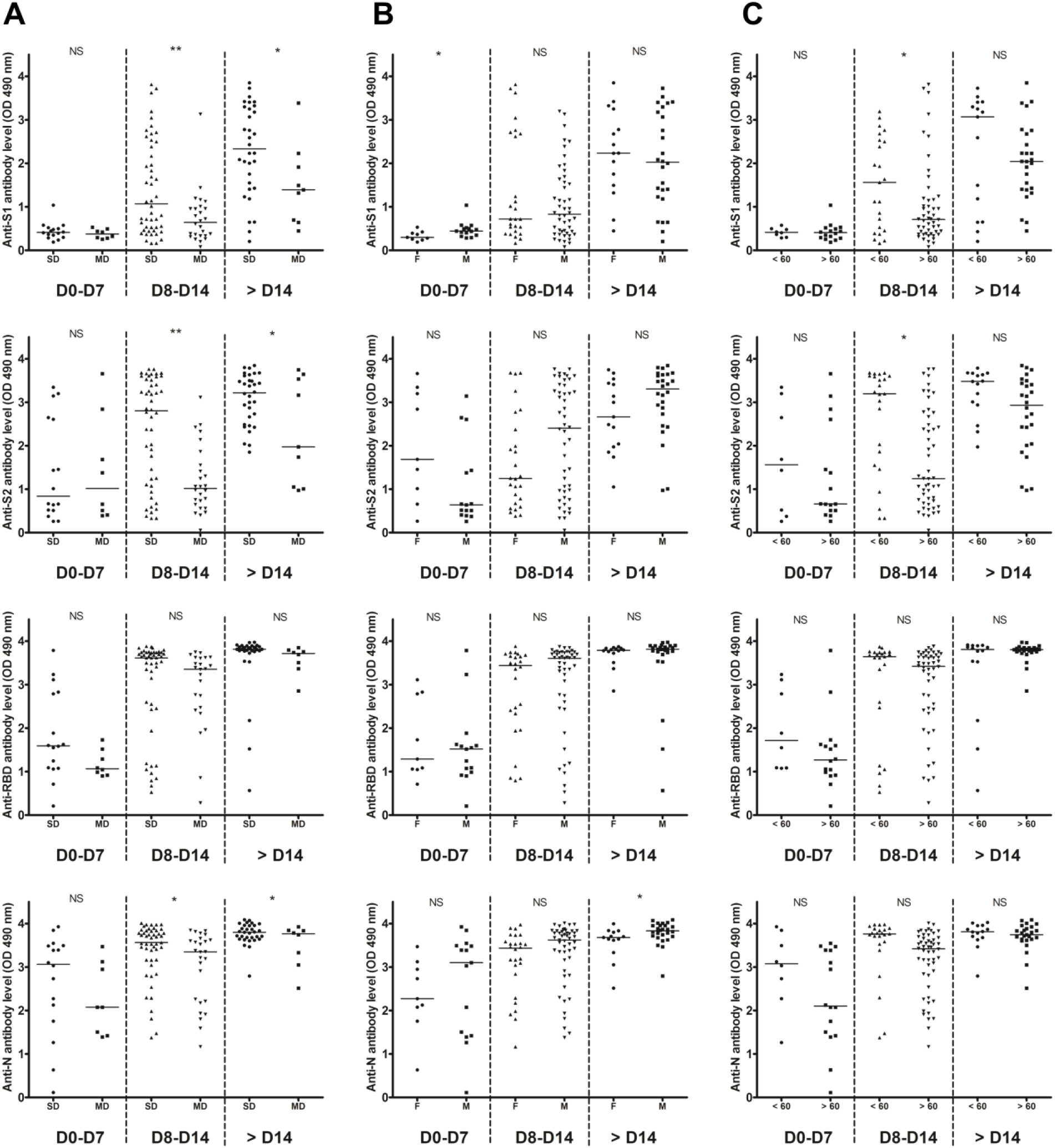
Temporal profiles of anti-S1, anti-S2, anti-RBD and anti-N antibody levels. Inpatients samples were divided into three periods groups (day 0-7, day 8-14 and day >14). (A) The temporal profiles are presented according to the severity of the disease (SD, severe disease requiring intensive care; MD, mild disease requiring non-intensive care). (B) The temporal profiles are presented according to the sex (M, male; F, female). (C) The temporal profiles are presented according to the age (< or > 60 years old). Dashed lines indicate assays cut-offs for positivity and lines indicate the median for each assay. OD, optical density. NS, not significant; *, p<0.05s; **, p<0.01.

**Supplementary Fig. 3.**
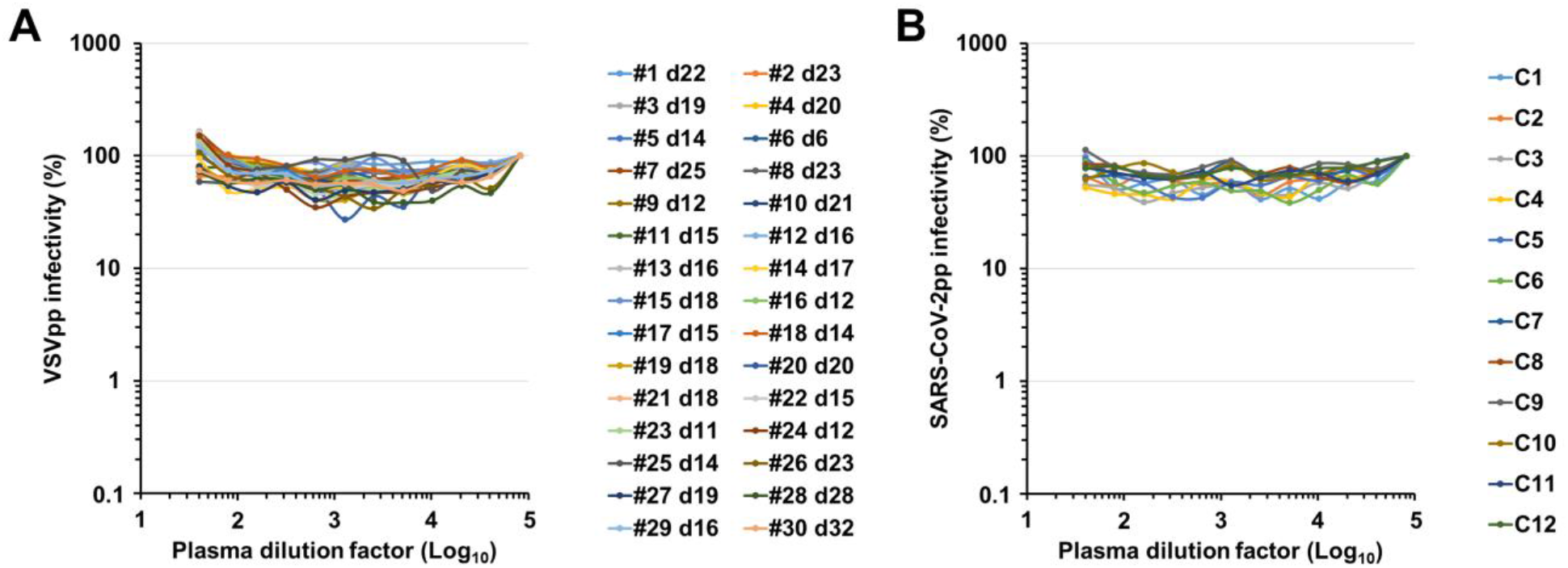
Specificity of the neutralization assay. (A) VSVpp were preincubated with serially diluted plasma obtained from 30 COVID-19 inpatients (#1 to #30). (B) SARS-CoV-2pp were preincubated with serially diluted plasma obtained from 12 patients infected with 229E, NL63, HKU1 or OC43 coronaviruses (C1 to C12). Dose response curves represent the means of normalized infectivity (%) from two independent experiments performed in duplicate. Error bars have been omitted for clarity.

**Supplementary Fig. 4.**
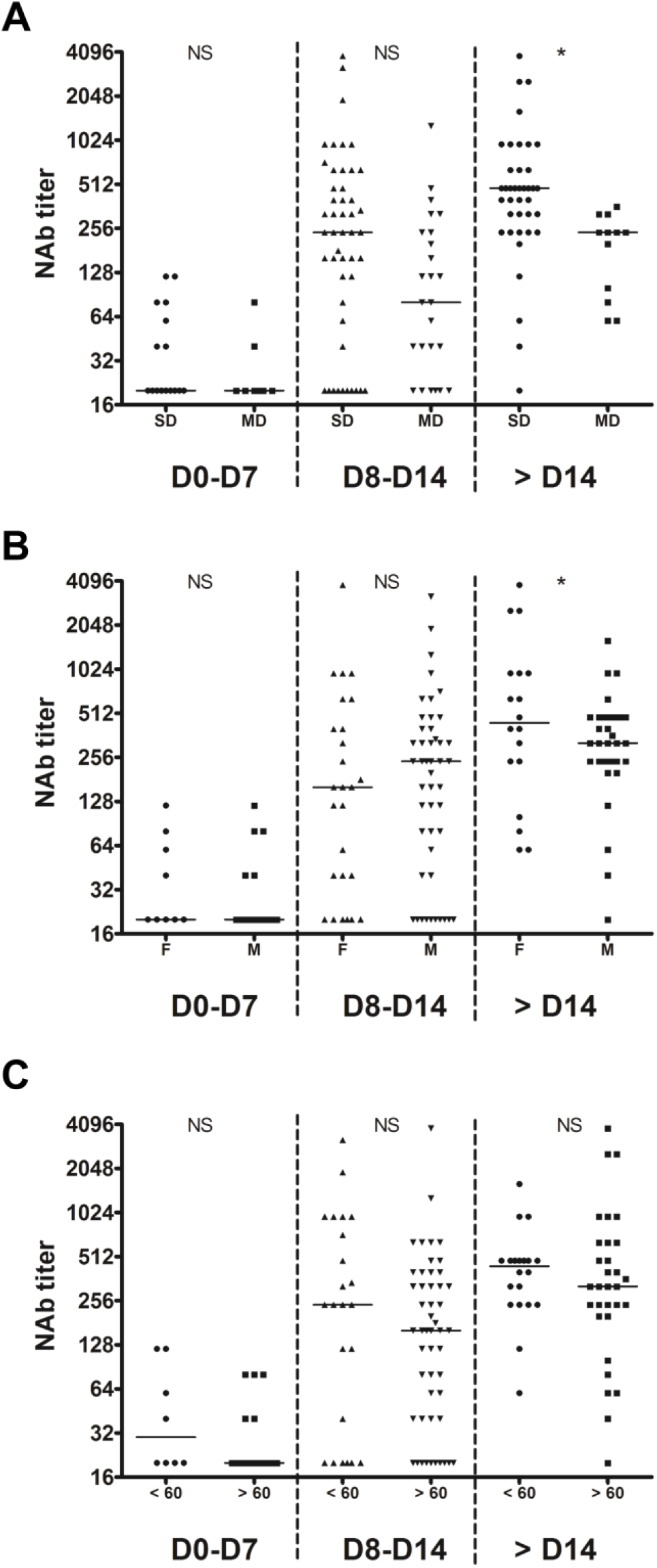
Temporal profiles of NAb titers. Inpatients samples were divided into three periods groups (day 0-7, day 8-14 and day >14). (A) The temporal profiles are presented according to the severity of the disease (SD, severe disease requiring intensive care; MD, mild disease requiring non-intensive care). (B) The temporal profiles are presented according to the sex (M, male; F, female). (C) The temporal profiles are presented according to the age (< or > 60 years old). Dashed lines indicate assays cut-offs for positivity and lines indicate the median for each assay. NS, not significant; *, p<0.05; **, p<0.01.

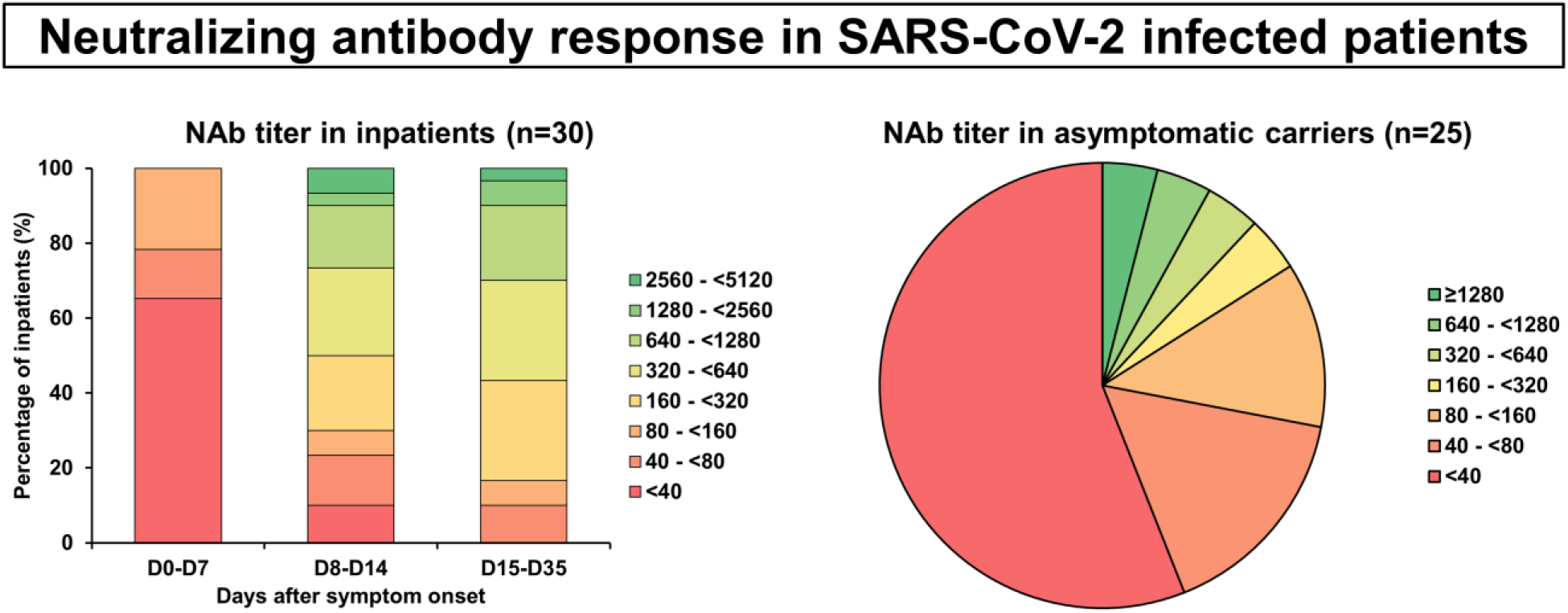

